# FDA orphan drug designations for lysosomal storage disorders – a cross sectional analysis

**DOI:** 10.1101/2020.01.05.20016568

**Authors:** Sven F. Garbade, Matthias Zielonka, Konstantin Mechler, Stefan Kölker, Georg F. Hoffmann, Christian Staufner, Eugen Mengel, Markus Ries

**Author notes:** **Correspondence to:** Markus Ries, MD, PhD, MHSc, FCP, Division of Pediatric Neurology and Metabolic Medicine, Center for Pediatric and Adolescent, Medicine, University Hospital Heidelberg, Im Neuenheimer Feld 430, D-69120 Heidelberg, Germany.

## Abstract

**Purpose:** To provide a quantitative clinical-regulatory insight into the status of FDA orphan drug designations for compounds intended to treat lysosomal storage disorders (LSD’s).

**Methods:** Assessment of the drug pipeline through analysis of the FDA database for orphan drug designations with descriptive and comparative statistics.

**Results:** Between 1983 and 2019, 124 orphan drug designations were granted by the FDA for compounds intended to treat 28 lysosomal storage diseases. Orphan drug designations focused on Gaucher disease (N=16), Pompe disease (N=16), Fabry disease (N=10), MPS II (N=10), MPS I (N=9), and MPS IIIA (N=9), and included enzyme replacement therapies, gene therapies, and small molecules, and others. Twenty-three orphan drugs were approved for the treatment of 11 LSDs. Gaucher disease (N=6), cystinosis (N=5), Pompe disease (N=3), and Fabry disease (N=2) had multiple approvals, CLN2, LAL-D, MPS I, II, IVA, VI, and VII one approval each. This is an increase of nine more approved drugs and four more treatable LSD’s (CLN2, MPS VII, LAL-D, and MPS IVA) since 2013. Mean time between orphan drug designation and FDA approval was 89.7 SD 55.00 (range 8-203, N=23) months.

**Conclusions:** The development pipeline is growing and evolving into diversified small molecules and gene therapy. CLN2 was the first and only LSD with an approved therapy directly targeted to the brain. Newly approved products included “me-too” – enzymes and innovative compounds such as the first pharmacological chaperone for the treatment of Fabry disease.

## Introduction

Lysosomal storage disorders (LSDs) are a group of more than 50 inherited, multisystemic, progressive conditions caused by a genetic defect that results in the progressive accumulation of complex non-metabolized substrates in the lysosomes of cells, tissues and organs, inducing distinct but heterogeneous somatic and neurological disease phenotypes [1-7]. In general, lysosomal storage disorders lead to significant morbidity and decreased life expectancy. Reported prevalences of LSDs in industrialized countries range between 7.6 per 100,000 live births (=1 in 13,158) and 25 per 100,000 live births (=1 in 4000) [8-11]. Some LSDs are treatable and the drug development in the field has traditionally been very active and dynamic after the successful development of enzyme replacement therapy in Gaucher disease which seeded further innovation [1, 12, 13]. The development of new compounds and new concepts of treatment for lysosomal storage disorders has been very dynamic. Therefore, the purpose of the present paper is to precisely analyze the most recent advances and to document novel trends in orphan drug development for lysosomal storage diseases as documented in the FDA Orphan Drug Product designation database.

## Methods

The FDA orphan drug database was accessed over the internet at the following address http://www.accessdata.fda.gov/scripts/opdlisting/oopd/. Search criteria were “all designations” from 1 January 1983 until 10 May 2019, i.e., all data entries until 10 May 2019 were taken into account (N=4979). The output format was an excel file which was downloaded on a local computer. Orphan designations for lysosomal storage diseases were extracted with pertinent keywords. (N=124) [1]. STROBE criteria were respected [14].

### Definitions

Pharmacological compounds were categorized based on their chemical structure into the following classes, listed in alphabetical order: “enzyme”, “enzyme/small molecule combination”, “gene therapy”, “polymer”, “protein (other than enzyme)”, and “small molecule” [1]. A small molecule was defined as a compound with a molecular weight below 900 Da [15]. In addition, compounds were further grouped into functionally meaningfully subtypes based on their biochemical properties, molecular mechanism of actions, or gene therapy platforms, i.e., (in alphabetical order): “AAV vector”, “adjunctive therapy”, “anaplerotic”, “anti-inflammatory/neuroprotective”, “anti-inflammatory/pro-chondrogenic”, “anti-inflammatory/TPP1 enhancing”, “anti-inflammatory/TPP1 enhancing/vitamin combination”, “pharmacological chaperone”, “cytochrom P450 rescue”, “enzyme replacement therapy”, “enzyme replacement therapy –pharmacological chaperone co-administration”, “lentiviral vector”, “membrane stabilization”, “lysosomal cholesterol redistributor”, “replacement therapy with a modified enzyme”, “nonviral vector directing transgene integration”, “receptor amplification”, “retroviral vector”, “small molecule facilitating intracellular substrate transport”, “stem cells”, “stop codon read-though”, “substrate reduction”.

Time to FDA approval was defined as the time period from orphan drug designation until approval by the FDA [1]. Drug approval rates were defined the proportion of orphan drug designations *approved* out of overall orphan drug designations *granted*. Missing data were not imputed.

### Statistical analysis

Standard techniques of descriptive statistics were applied: continuous variables were summarized with mean, standard deviation, median, minimum and maximum values. Categorical variables were summarized with frequencies and percentages.

Comparative statistics were performed with the appropriate parametric test for data with Gaussian distribution (i.e., ANOVA). Differences between frequency counts were compared with the chi-square test. Two-sided p-value□<□0.05 was considered statistically significant All statistical analyses were performed using SAS Enterprise guide 7.13 HF4, SAS Institute Inc., Cary, NC, USA. Graphs were generated with R [16] and GraphPad Prism 5.04, GraphPad Software, Inc., San Diego, CA, USA.

## Results

The drug development pipeline: orphan drug designations granted by the FDA Between 1 January 1983 and 10 May 2019, 124 orphan drug designations were granted by the FDA for compounds intended to treat 28 lysosomal storage diseases (Figure 1A). For a comparison of dimensions, in the same time period, the FDA granted 4979 orphan drug designations overall, out of which 783 were approved (Figure 1B). Twenty conditions had multiple orphan drug designations. Most orphan drug designations were granted for Gaucher disease (N=16), Pompe disease (N=16), Fabry disease (N=10), MPS II (N=10), MPS I (N=9), and MPS IIIA (N=9), followed by 14 others depicted in Figure 2A. Eight conditions had one orphan drug designation. Enzyme replacement therapies, gene therapies, small molecules, and other technology platform classes were designated as orphan drugs intended to treat lysosomal storage diseases (Figure 2B). Nine granted orphan drug designations were subsequently withdrawn (Table 1). The reason for withdrawal is not specified in the FDA orphan drug database.

**Table 1:**
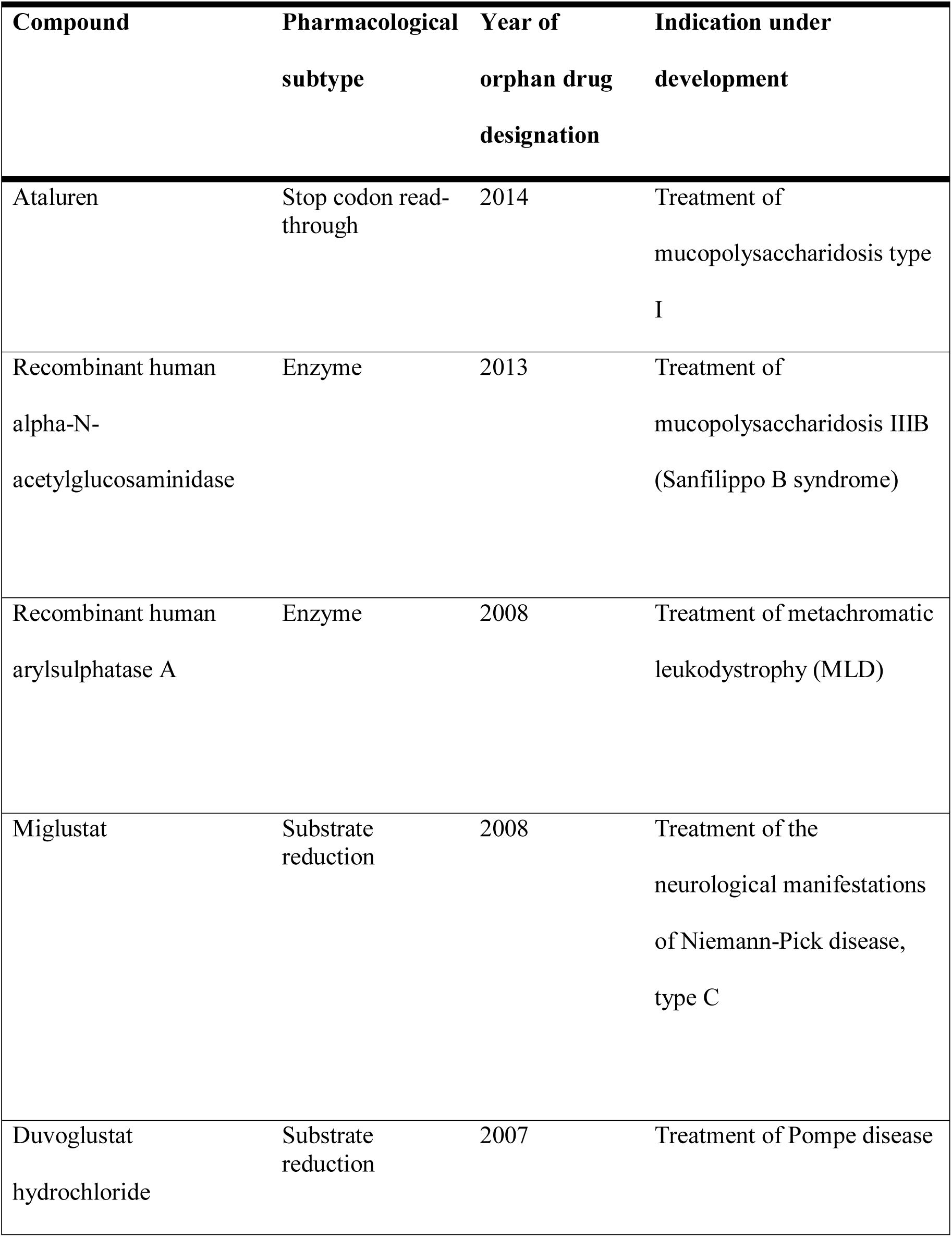
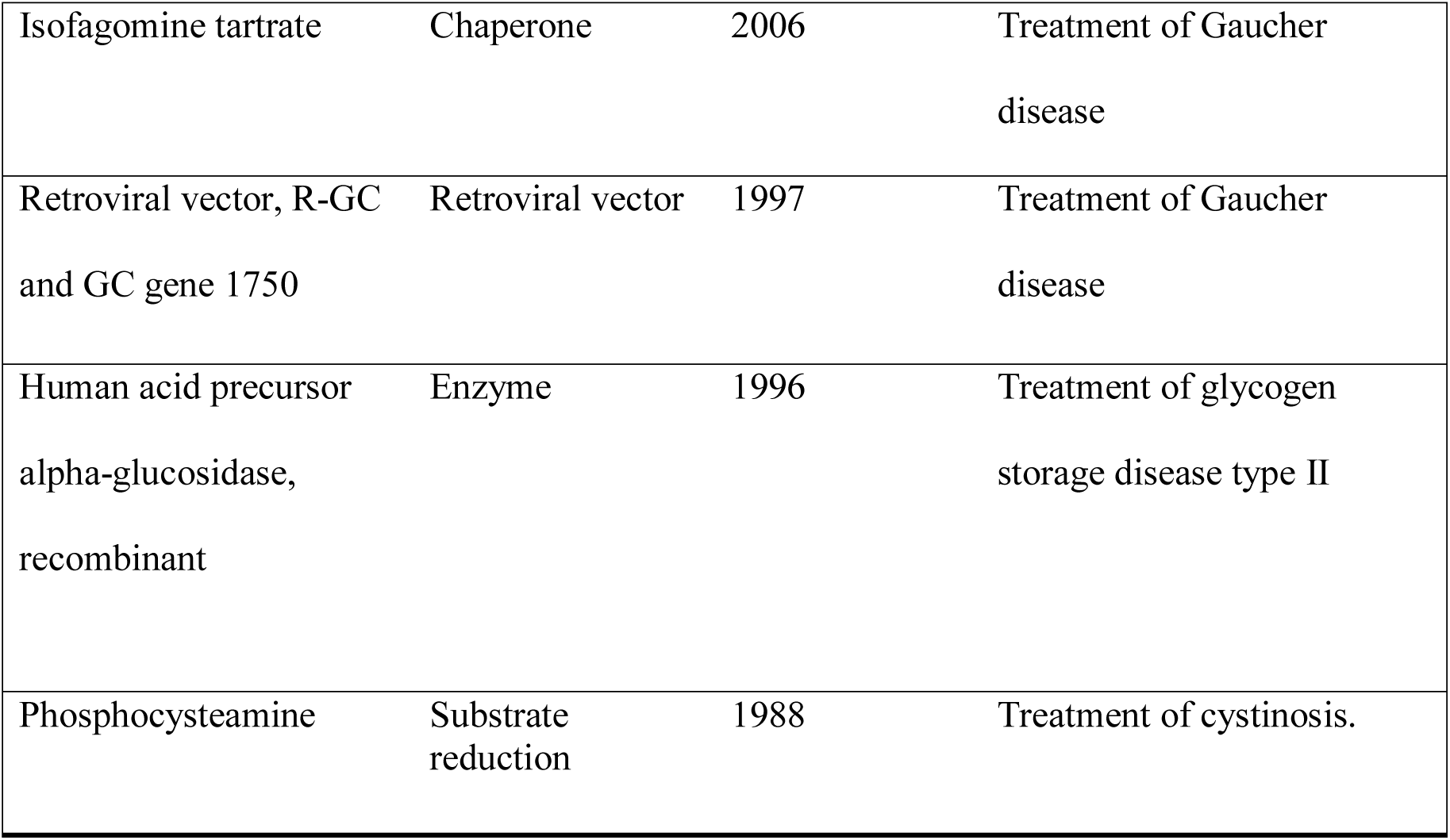
Withdrawn orphan drug designations. Reasons for and time of withdrawal were not specified in the FDA database

| **Compound** | **Pharmacological subtype** | **Year of orphan drug designation** | **Indication under development** |
| --- | --- | --- | --- |
| Ataluren | Stop codon read-through | 2014 | Treatment of mucopolysaccharidosis type I |
| Recombinant human alpha-N-acetylglucosaminidase | Enzyme | 2013 | Treatment of mucopolysaccharidosis IIIB (Sanfilippo B syndrome) |
| Recombinant human arylsulphatase A | Enzyme | 2008 | Treatment of metachromatic leukodystrophy (MLD) |
| Miglustat | Substrate reduction | 2008 | Treatment of the neurological manifestations of Niemann-Pick disease, type C |
| Duvoglustat hydrochloride | Substrate reduction | 2007 | Treatment of Pompe disease |
| Isofagomine tartrate | Chaperone | 2006 | Treatment of Gaucher disease |
| Retroviral vector, R-GC and GC gene 1750 | Retroviral vector | 1997 | Treatment of Gaucher disease |
| Human acid precursor alpha-glucosidase, recombinant | Enzyme | 1996 | Treatment of glycogen storage disease type II |
| Phosphocysteamine | Substrate reduction | 1988 | Treatment of cystinosis. |

**Figure 1A:**
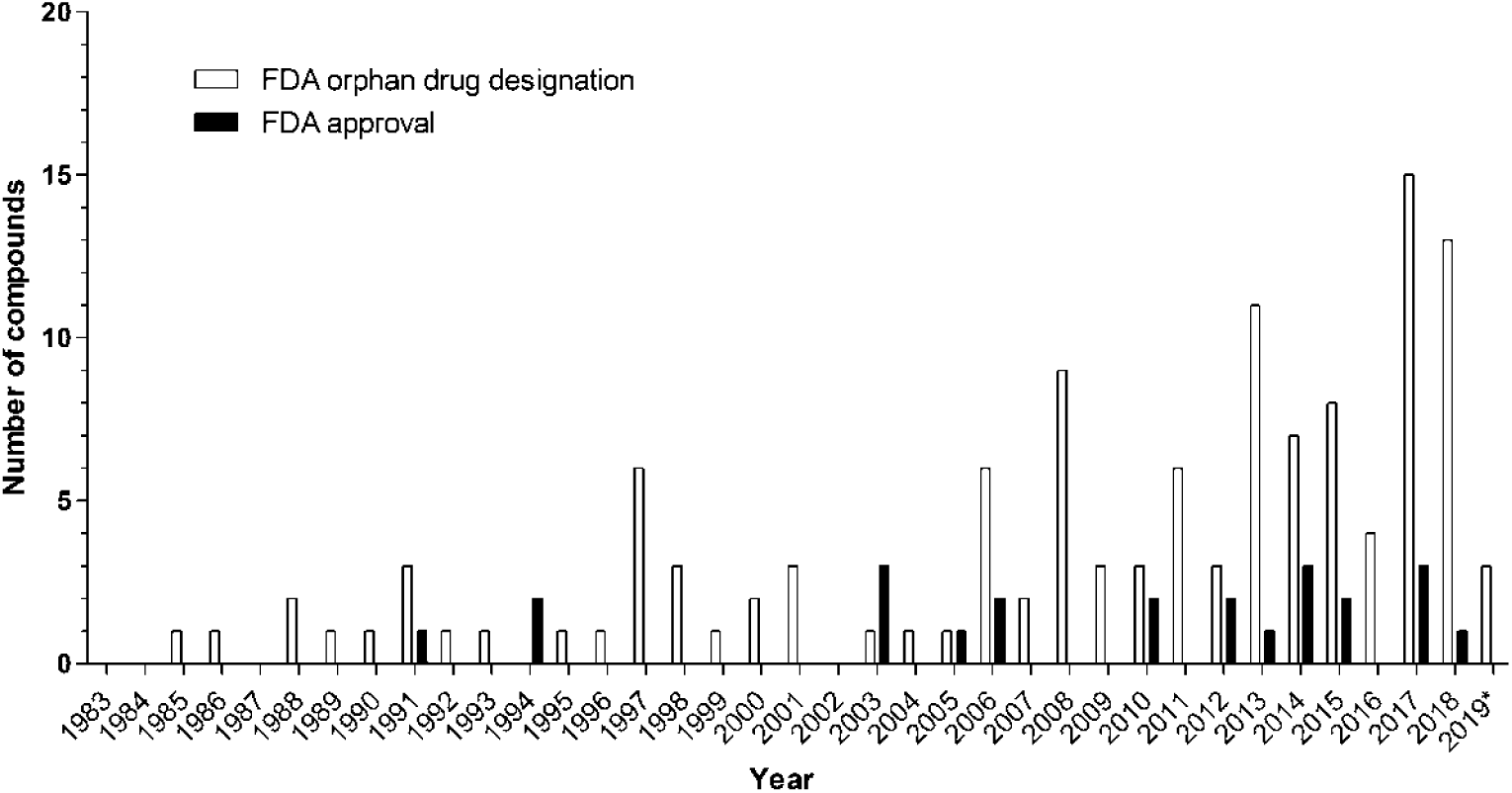
Number of orphan drug designations (open bars) and FDA approvals (full bars) for compounds intended to treat lysosomal storage diseases by year. * indicates close of database: 10 May 2019

**Figure 1B:**
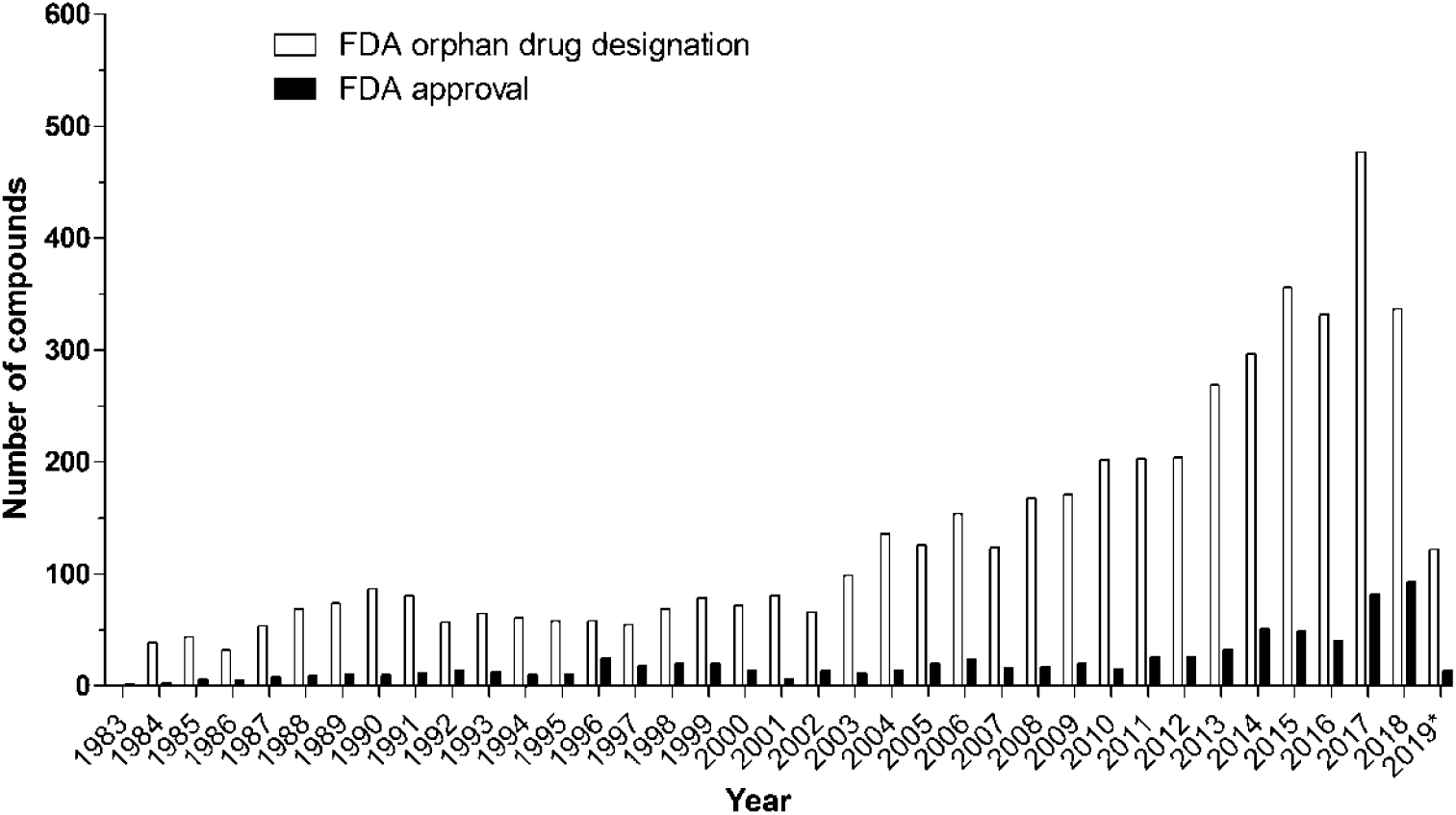
Overall number of orphan drug designations (open bars) and FDA approvals (full bars) by year. *indicates close of database: 10 May 2019

**Figure 2A:**
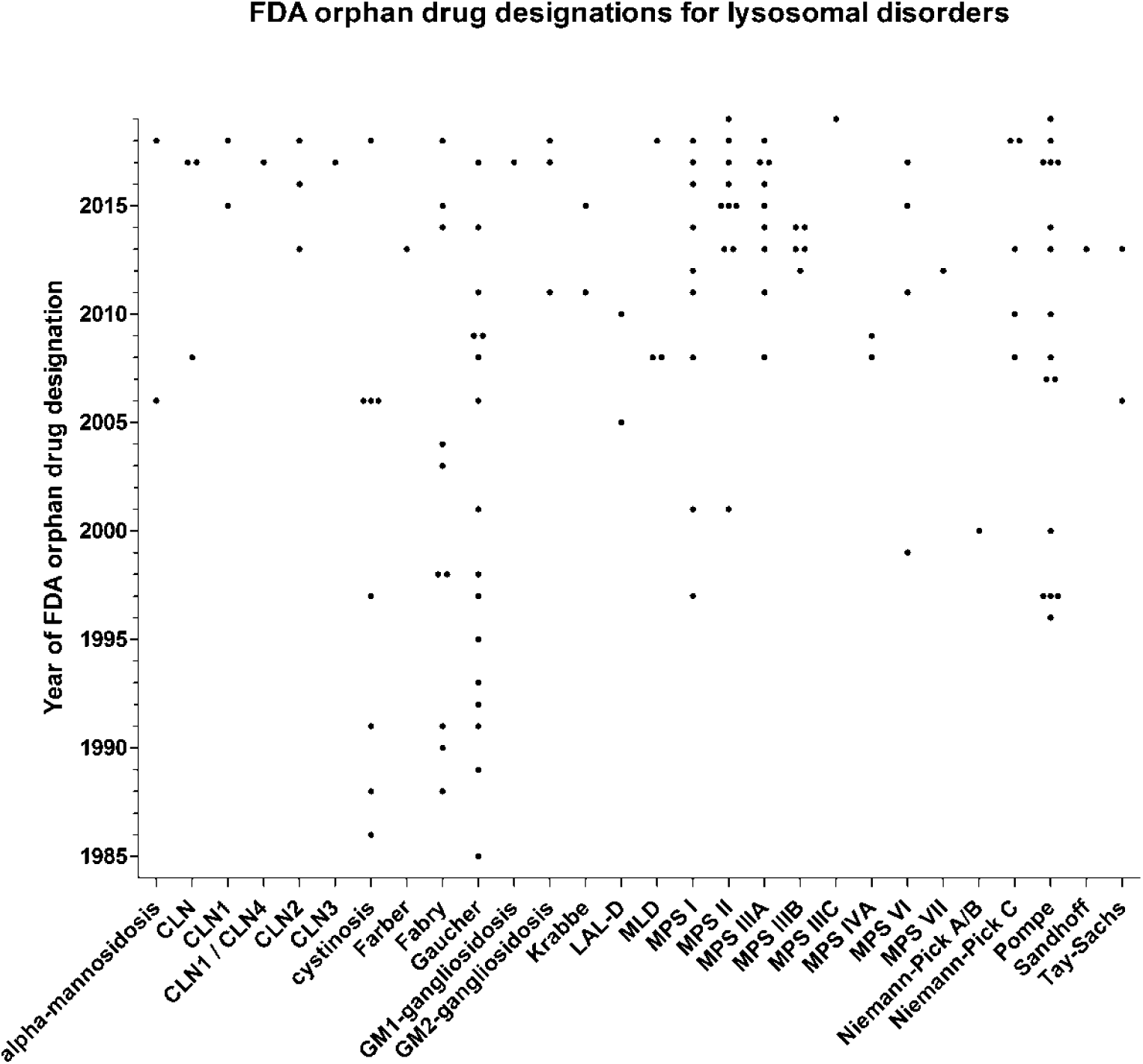
Orphan drug designations granted by the FDA for compounds intended to treat lysosomal storage disorders by year and specific disease.

**Figure 2B:**
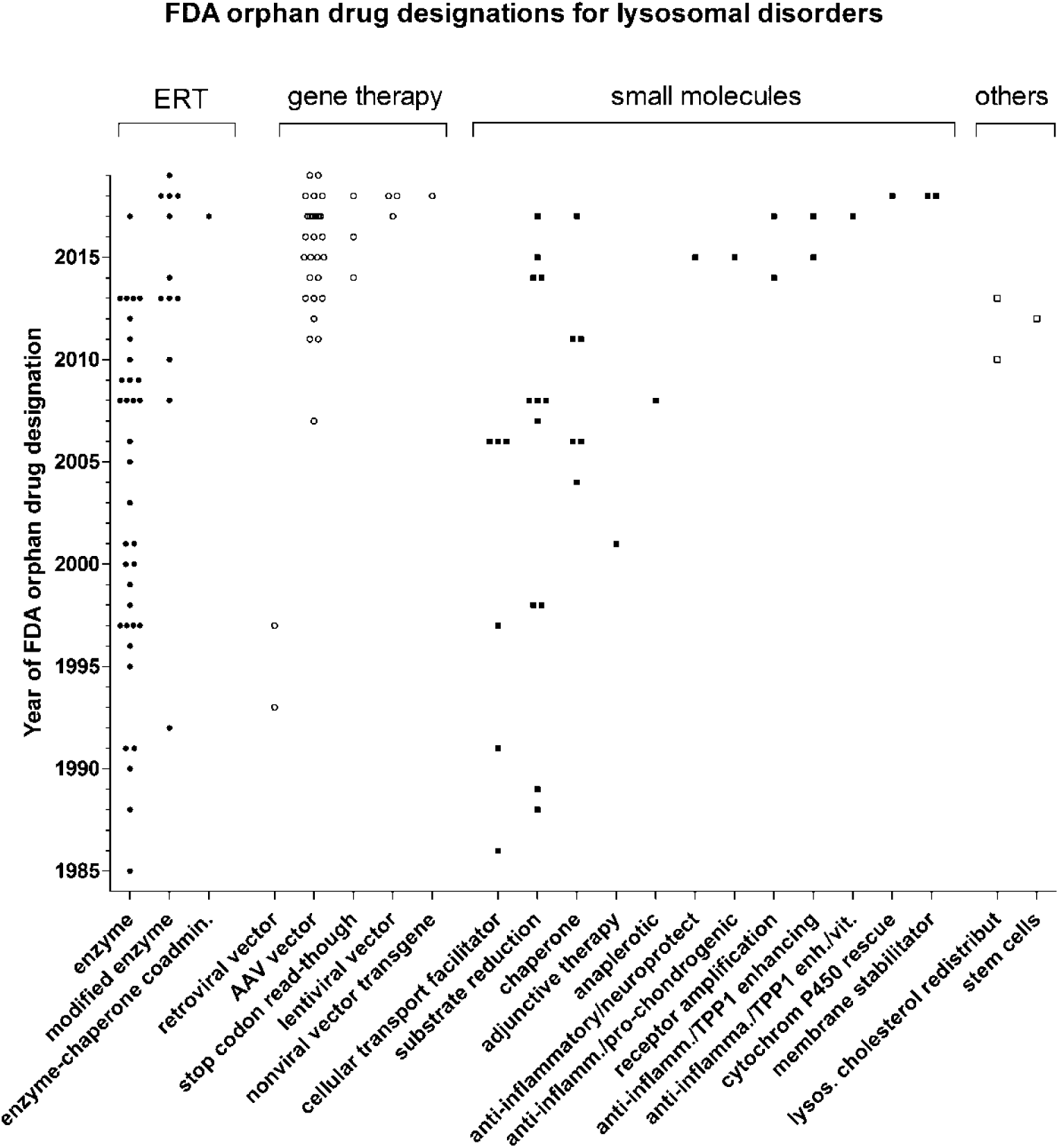
Orphan drug designations granted by the FDA for compounds intended to treat lysosomal storage disorders by year and pharmacological technology platform.

The approval rate of *lysosomal* orphan drugs (18.5%) did not differ from approval rates for *non-lysosomal* orphan drugs (15.7%, p=0.38, chi-square)

### Lysosomal storage disorders with FDA approved therapies

Twenty-three orphan drugs were approved for the treatment of 11 lysosomal storage diseases. Four diseases had multiple therapeutics approved, i.e. Gaucher disease (N=6), cystinosis (N=5), Pompe disease (N=3), and Fabry disease (N=2), (Figure 3A). The remaining seven diseases had one compound each approved by the FDA (i.e., CLN2, LAL-D, MPS I, II, IVA, VI, VII). CLN2 was the only neuronopathic lysosomal storage disease with an FDA approved therapy directly targeting the brain; all the other therapies address systemic non-neurological manifestations. FDA approved therapies included enzyme replacement therapies (N=15) and small molecules (N=8), but no other class of drugs (Figure 3B, Table 2).

**Table 2.**
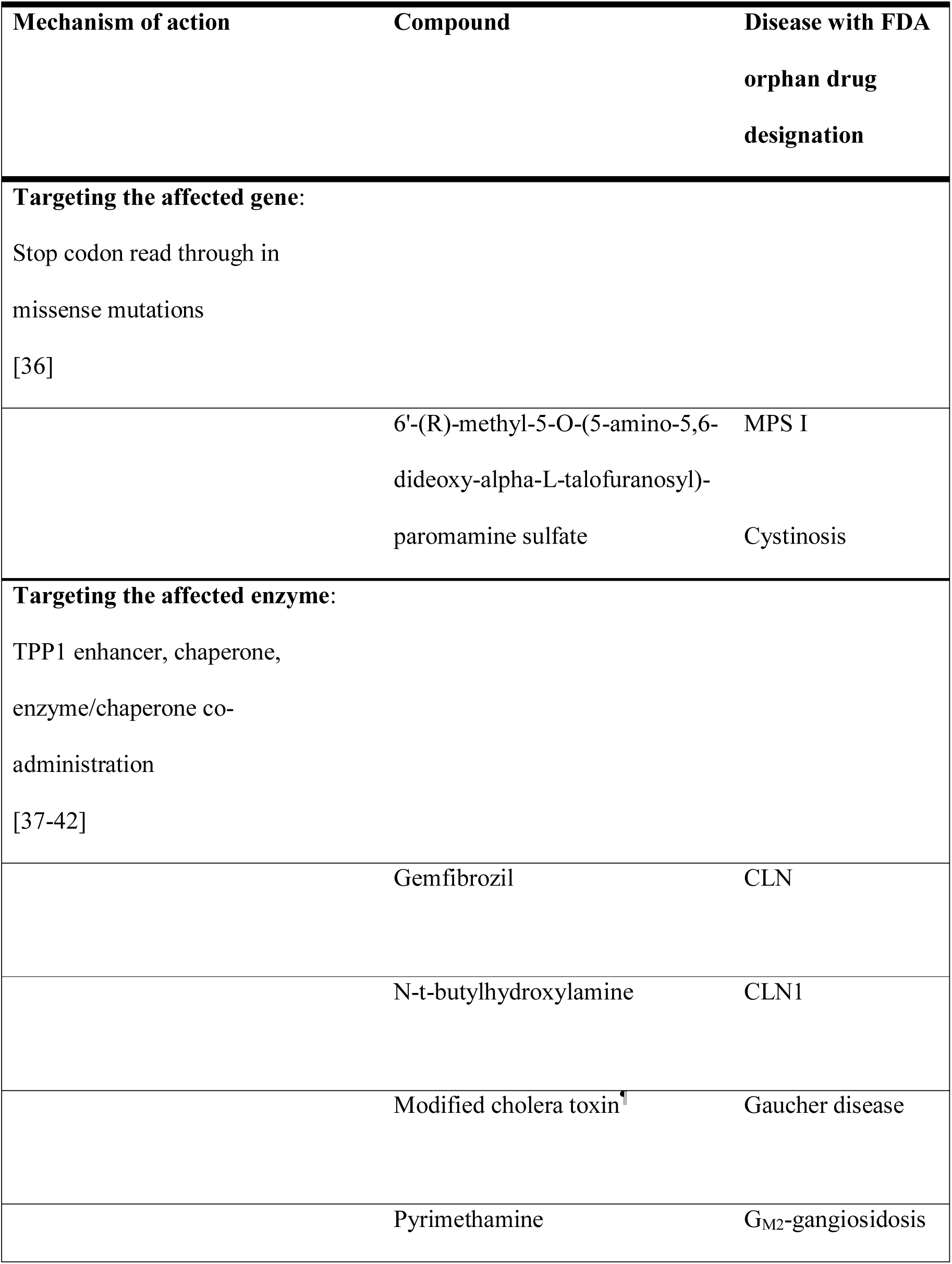
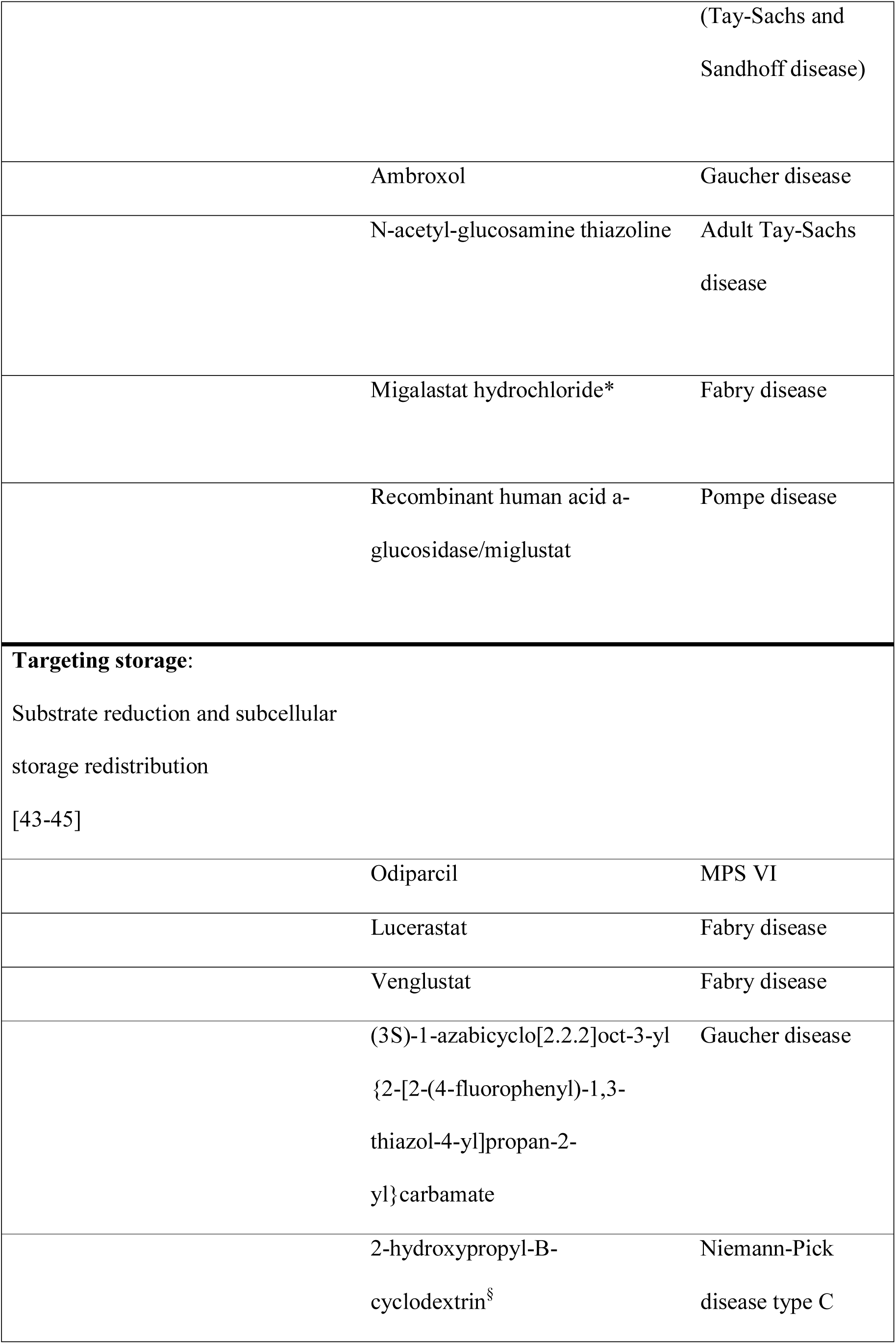
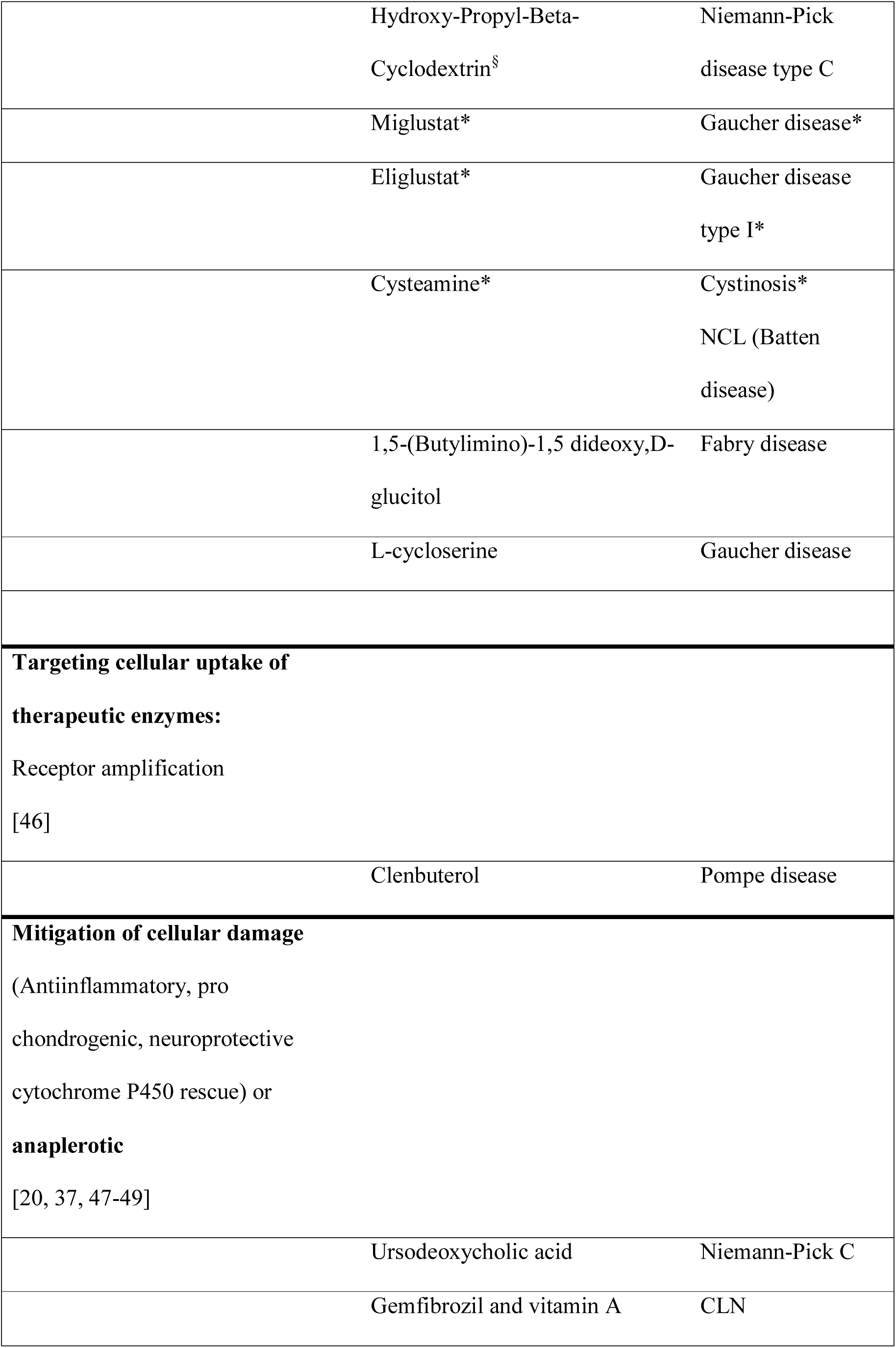
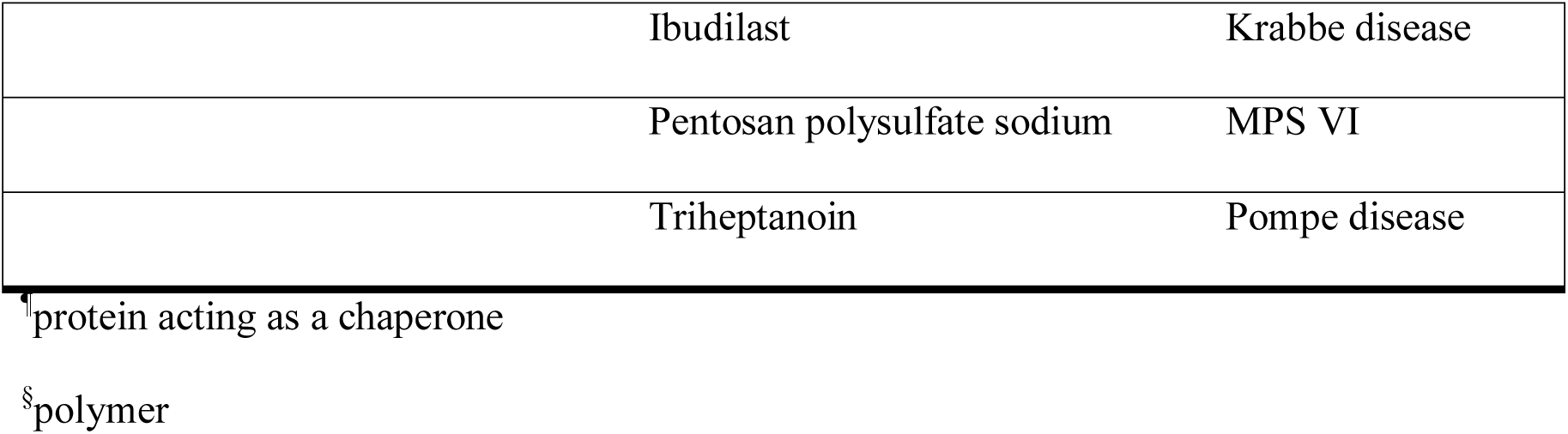
Mechanism of action of FDA approved small molecules (*) and small molecules in development, intended to treat a lysosomal storage disorder.

**Figure 3A:**
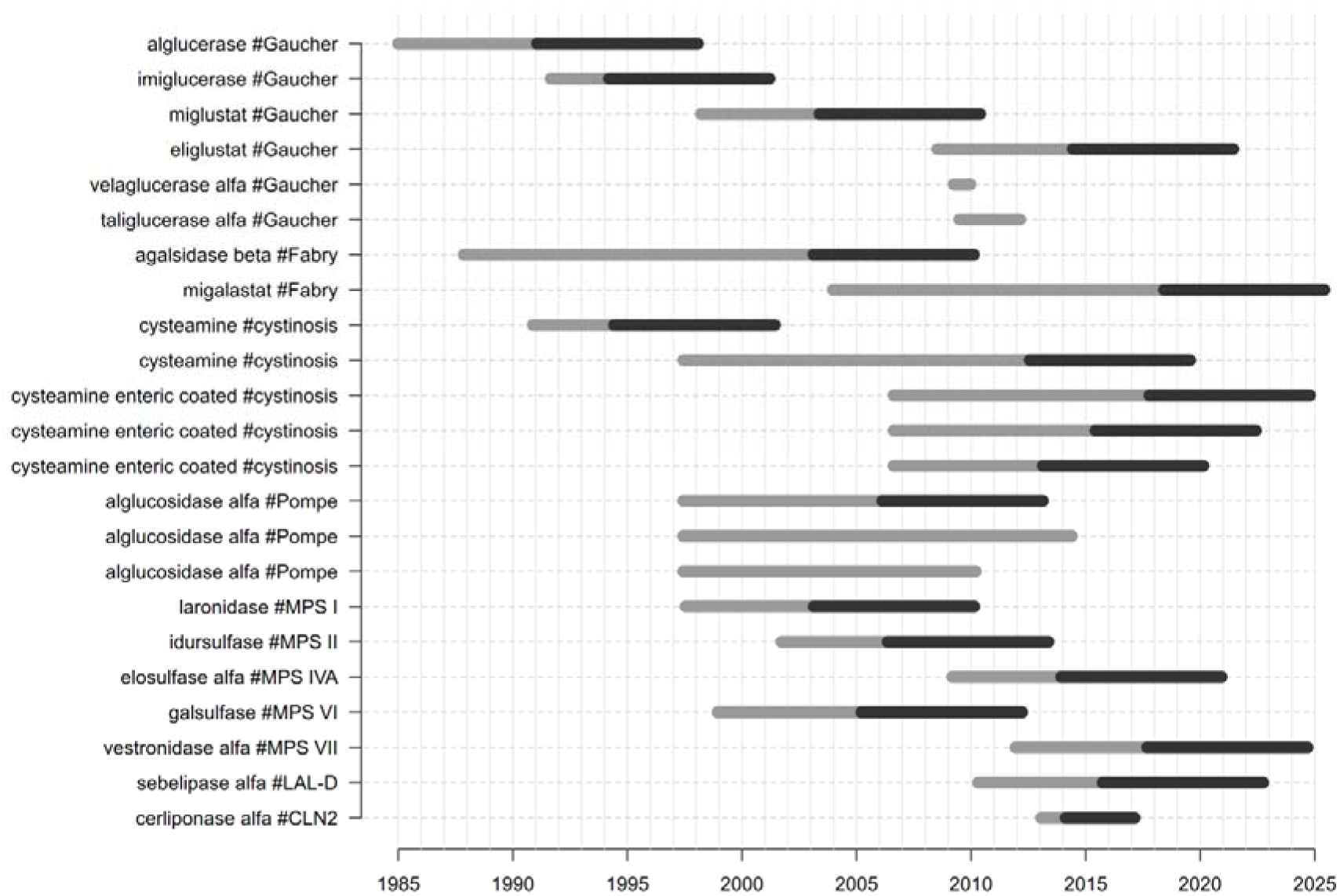
FDA approved compounds for the treatment of lysosomal storage disorders (depicted as compound#disease), development times and market exclusivity. Grey bars indicate drug development times, i.e. time from orphan drug designation to orphan drug approval by the FDA. Black bars indicate, if applicable, market exclusivity periods. Treatments for cystinosis included three different formulations (systemic, systemic extended release, and ophthalmic), and three different age groups (adults, children 2-6 years, and children 1 year of age to less than 2 years of age.

**Figure 3B:**
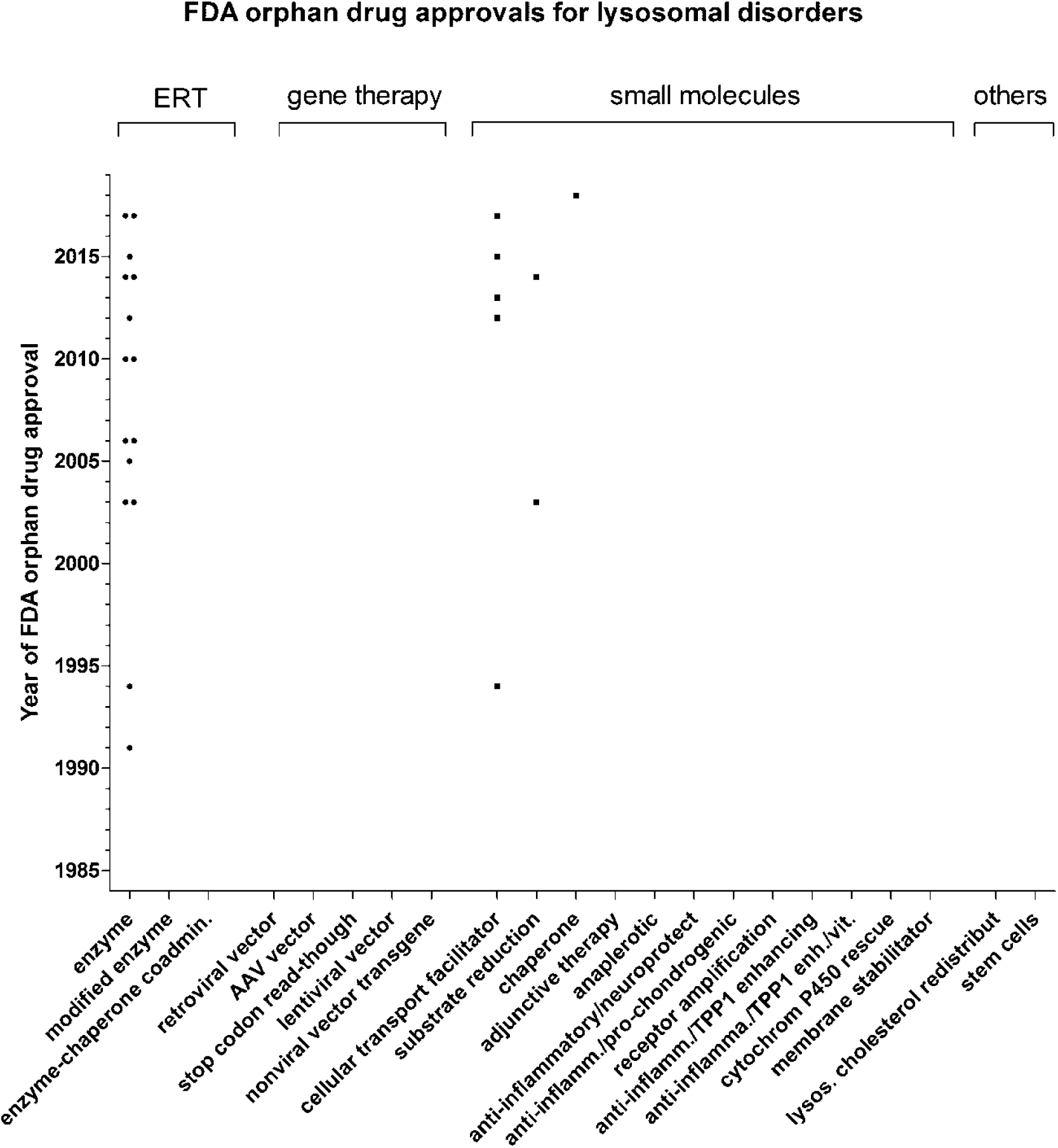
FDA approved therapies for the treatment of lysosomal storage disorders by year of approval and pharmacological technology platform

### Drug development timelines

Overall mean time to approval, defined as time between orphan drug designation and FDA approval was 89.7 SD 55.00 (range 8-203, N=23) months. Stratified by drug compound subtypes, mean time to approval for enzyme replacement therapies was 81.2 SD 56.42 (range 8-203, N=15) months, mean time to approval for small molecules facilitating subcellular transport was 107.8 SD 52,96 (range 40-181, N=5) months, and mean time to approval for substrate reduction therapies was 66.5 SD 6.36 (range 62 to 71, N=2) months. Time to approval for the pharmacological chaperone therapy was 173 months. Differences between the groups were not statistically significant (p=0.33, ANOVA). The drug development timelines and market exclusivity periods, an incentive granted by the FDA to stimulate orphan drug development [13], are illustrated in Figure 3A.

## Discussion

By 10 May 2019, 23 orphan drugs were approved by the FDA for the treatment of 11 lysosomal storage disorders. This is an increase of nine more approved orphan drugs and four more treatable lysosomal disease (i.e. CLN2, MPS VII, LAL-D, and MPS IVA) compared to 2013 [1].

While alglucerase for Gaucher disease was the first orphan drug approved for a lysosomal storage disease in 1991, intrathecally administered cerliponase alfa for CLN2, FDA approved in 2017, is the first orphan drug approved to directly treat the brain which is a significant therapeutic innovation [17, 18]. Since 2013, 54 more orphan drug designations were granted. In addition, diseases such as CLN1, CLN3, CLN4, Farber disease, and G_M1_-gangliosidosis did not have orphan drug designations in 2013, which indicates that drug development in lysosomal storage disorders is now being driven into mainly neuronopathic conditions (Figure 2A). The overall growth curve of orphan drug designations appears to accelerate over time and may become exponential (Figure 1A), which may follow indeed a global trend (Figure 1B). Of interest, the drug approval rate in lysosomal orphan drug development and non-lysosomal orphan drug development did not differ. Technology is evolving: while enzyme replacement therapies had initially set the trend, more modified enzymes, including fusion-proteins, and an enzyme-chaperone co-administration entered the development pipeline. This may be a reaction to the increasing recognition in the field, that, in general, systemically administered enzyme replacement therapy with conventional enzymes can easily access organs such as liver and spleen, but have little impact on bone and CNS manifestations. Four small molecules have been approved by the FDA for the treatment of a lysosomal storage disease. Their mechanisms of action target the facilitation of subcellular transport (e.g., cysteamine for cystinosis, approved in 1994), and the reduction of storage (miglustat, approved in 2003, and eligustat, approved in 2014, both for the treatment of Gaucher disease) [1]. In 2018, migalastat, which stabilizes the misfolded enzyme alpha-glactosidase A, was approved as first-of-its kind pharmacological chaperone by the FDA for the treatment of Fabry disease [19], (Table 2). Mechanisms of action for small molecules, either approved or in drug development, considered the broad spectrum of underlying pathophysiology and aimed at 1) targeting the affected gene 2) targeting the affected enzyme 3) targeting storage 4) targeting cellular uptake of therapeutic enzymes, and 4) mitigation of cellular damage or anaplerotic (Table 2). It is possible and likely that not all mechanistically meaningful approaches lead to clinical benefit in patients [20]. The plethora of innovative ideas for pharmacological approaches is laudable, but should not lead a treating physician to engage in off-label use, but rather encourage international collaboration aimed to generate the highest standard of evidence-based knowledge by respecting excellence clinical research [21]. Gene therapy now plays a larger role in the drug development pipeline compared to the situation in our last analysis [22]. This may again be a reaction to the increased recognition of enzyme replacement therapies’ substantial limitations, as described in detail above. The technical approach towards gene therapy is evolving as illustrated in Figure 2B. Gene therapies rely at least in principle on the assumption that a single treatment may result in a sustained, potentially curative clinical benefit for the patients. The first molecular tools enabling efficient non-toxic gene transfer into human somatic cells were recombinant replication-deficient vectors [23]. Among those, retroviral and adeno-associated viral (AAV) vectors have been the most widely used in particular for *ex vivo* T cell engineering or genetically engineered hematopoietic stem cells (HSCs) for the treatment of primarily hematologic or oncologic conditions such as pediatric ALL, β-thalassemia or adenosine deaminase deficiency [24-26]. In contrast, while the first two orphan drug designations for gene therapy for lysosomal storage diseases in 1993 and 1997 (both for Gaucher disease) relied on retroviral vectors, this platform was subsequently abandoned. This is likely due to the emergence of serious toxicities related to high gene transfer including insertional genotoxicity, immune destruction of genetically modified cells, and immune reactions related to the application of certain vectors [27, 28]. The next step in gene technology was the introduction of AAV (designated for Pompe disease in 2007), followed by stop-codon read through (designated in 2014 and 2016 for MPS I, and in 2018 for cystinosis). Moreover, lentiviral vector (designated in 2018 for Fabry disease and MLD), and nonviral vector directing transgene integration (designated in 2018 for MPS I) technologies are being considered, all of which have to prove their safety and efficacy in the future. More sophisticated genome editing technologies that enable a variety of therapeutic genome modifications (gene addition, gene ablation or “gene correction”) consist of the administration of transcription activator-like effector nucleases (TALENs) and or CRISPR-Cas 9 system to efficiently cleave and modify DNA at sites of interest [29-33]. Those approaches are currently limited to applications in basic research, but transfer into clinical trials can be expected in the near future [34, 35]. Until close of database no gene therapy was approved for the treatment of lysosomal storage disorders (Figure 3B). If proven successful in registration trials - which would supposedly be small clinical trials of a limited duration - it is of particular interest, how long the therapeutic effect of gene therapy can be sustained during a patients’ lifetime, and, if this time is limited, whether it would be safe and feasible to repeat the administration of a gene therapy multiple times in a single patient. It is anticipated that novel therapies will be costly. Important topics for future investigations will include patient selection, starting and stopping criteria.

For the correct contextual interpretation it is important to be aware of important limitations of the present work as pointed out previously [1]. Orphan drug designation was considered as the expressed intent to develop a drug. This may be biased by strategic and patent related considerations and not all manufacturers may choose to go this publically visible pathway from early on. Time of orphan drug designation may be somewhat arbitrary in the drug development process; therefore, time-to-approval as presented in the present analysis may also be biased by the intellectual property strategy of the respective drug development program. Orphan drug development outputs in jurisdictions other than the FDA were, similar to our previous analysis, not taken into account because this analysis was by definition focused on the impact of the US orphan drug act [1, 13]. As drug development in lysosomal storage disorders is, in general, a global enterprise we consider the present finding within the context of their limitations generalizable.

## Conclusions

Activities in orphan drug development for lysosomal storage disorders are steadily increasing which follows a global trend in orphan drug development overall. Newly approved products included “me-too” – enzymes, but also innovative compounds such as the first ERT targeting the brain in CLN2 and the first-of-its-kind pharmacological chaperone for the treatment of Fabry disease. The development pipeline is increasingly evolving into diversified small molecules and, in particular, gene therapies.

## Data Availability

all data generated or analysed during this study are included in this published article

## Declarations

### Ethics approval and consent to participate

not applicable

### Consent for publication

not applicable

### Availability of data and material

all data generated or analysed during this study are included in this published article

### Authors’ contributions

Substantial contributions to the conception or design of the work and supervision: MR Data acquisition, analysis, interpretation of data: SG, MZ, KM, SK, GFH, CS, EM, MR Drafting the work: MR

Substantively revision of the work: SG, MZ, KM, SK, GFH, CS, EM, MR

All authors have approved the submitted version (and any substantially modified version that involves the author’s contribution to the study).

All authors have agreed both to be personally accountable for the author’s own contributions and to ensure that questions related to the accuracy or integrity of any part of the work, even ones in which the author was not personally involved, are appropriately investigated, resolved, and the resolution documented in the literature.

## Acknowledgements

Not applicable

